# Beyond billing codes: a proof-of-concept study of large language models for pregnancy episode reconstruction from clinical notes

**DOI:** 10.64898/2026.11.19.26368521

**Authors:** Ann-Kathrin Schalkamp, Shadera Azzam, Aryan Ayati, Artur Belov, Michelle Wang, Vivek Rudrapatna

## Abstract

**Background:** Accurate identification of pregnancy episodes from electronic health records (EHRs) is essential for maternal and fetal health research. Existing approaches rely primarily on diagnosis and procedure codes, which may miss pregnancies documented outside the health system and are susceptible to coding errors. Clinical notes contain richer information but remain largely unexplored for this task.

**Objectives:** To evaluate whether a large language model (LLM) can reconstruct complete pregnancy episodes directly from clinical notes.

**Methods:** Chronologically concatenated notes and messages were provided to GPT-5, which identified episodes and extracted variables including assisted reproductive treatment, gestational age, outcome, and complications. Performance was benchmarked against manual chart review (gold standard) and a hierarchical ICD-code algorithm in 30 patients comprising 74 pregnancies (20 patients in a held-out test set).

**Results:** The LLM identified 53 of 54 test-set pregnancies versus 40/54 for a hierarchical ICD-code algorithm, with fewer spurious episodes (3 vs 24), corresponding to an F1 score for pregnancy detection against chart review of 0.96 vs 0.68. Among pregnancies identified by both methods, median absolute error was 0 vs 25.5 days for start dates (p<0.001) and 0 vs 1 day for end dates (p=0.044), and outcome accuracy was numerically but not significantly higher (0.84 vs 0.74 for exact outcome types; 0.92 vs 0.84 with abortion subtypes grouped). It additionally recovered information unavailable in structured data, including reproductive-treatment type and fetus-level outcomes.

**Conclusions:** An LLM applied to clinical narratives identified pregnancy episodes more completely and with more accurate dates than an ICD-code algorithm while capturing richer clinical detail. Because the two approaches used different input data, these gains reflect both the information in clinical notes and the LLM itself. LLM-based extraction from clinical notes is a promising complement to code-based pregnancy phenotyping that warrants evaluation in larger, multi-institutional cohorts.

## 1. Background and Significance

Pregnancy is a unique and complex part of the human lifecycle, creating specific challenges for epidemiological research and efforts to improve maternal and fetal outcomes^1,2^. Registries and trials remain the gold standard, but they are expensive, slow to build, narrow in scope, and often underpowered. Electronic health records (EHR) data are increasingly being used for pregnancy-related research, especially structured data elements like diagnosis and procedure codes^1,3–9^, but exhibit many key limitations: imprecision in estimating pregnancy start dates and thus gestational age, reduced performance for multiple gestations, and non-live birth outcomes, poor capture of out-of-system pregnancy history, sensitivity to coding practices and transitions, and limited portability across databases or institutions^1,3–9^.

Clinical notes contain substantially richer pregnancy-related information, including last menstrual period, gestational age assessments, ultrasound findings, evolving complications, and narrative descriptions of outcomes and care decisions. Further, historical pregnancies reported to the clinician will not be found in structured EHR data but can be documented in free text, for example with the abbreviation GTPAL (gravida, term, preterm, abortion, living). Traditional natural language processing (NLP) efforts in obstetrics have largely focused on identifying specific conditions or behaviors during pregnancy^10,11^, rather than reconstructing complete pregnancy episodes with explicit temporal boundaries from longitudinal clinical text. This is a critical gap, because many research questions require complete episodes to place exposures within pregnancy, link them to outcomes, and distinguish multiple pregnancies in the same person. To date, there is no widely adopted, validated approach that uses clinical narratives to derive pregnancy episodes at scale.

Large language models (LLMs) offer an opportunity to address these limitations. Unlike rule-based or code-centric approaches, LLMs can infer timelines directly from clinician documentation, reconcile conflicting signals, and extract clinically meaningful attributes such as complications and outcomes in a unified framework. LLMs can accurately ascertain pregnancy-related complications like postpartum hemorrhage^12^ as well as other clinical variables unrelated to pregnancy such as diagnoses, pathological classification^13^, patient reported outcomes^14^, and clinical disease scales^15^. LLMs have further successfully been applied for clinical temporal relation extraction^16–18^. However, these approaches largely use single notes in isolation, whereas pregnancy episode reconstruction requires consolidating fragmented and sometimes conflicting evidence across many encounters.

## 2. Objectives

Here we develop and evaluate an LLM-based method to characterize pregnancy episodes from unstructured clinical notes and patient messages at a single tertiary health care system (University of California, San Francisco [UCSF]). We compare its performance to manual chart-review, the gold-standard, as well as to a rule-based algorithm using International Classification of Diseases (ICD) codes.

## 3. Methods

### 3.1. Data

We conducted a retrospective study nested within an existing cohort of patients with inflammatory bowel disease. Using EHR data, we identified female patients and retrieved their ICD diagnosis and procedure codes related to pregnancy, delivery, postpartum care (Supplemental Table 1), as well as unstructured clinical notes from obstetrics and gynecology (OBGYN) providers, messages between patients and providers sent through the patient portal and messages between staff that discussed pregnancy (Supplemental Material). The study was approved by the University of California, San Francisco Institutional Review Board (Study #24-43008).

### 3.2. ICD-code algorithm

First, we implemented a hierarchical, rule-based algorithm (Supplemental Material, Supplemental Figure 1) to identify and characterize discrete pregnancy episodes from longitudinal ICD codes. ICD codes were mapped to pregnancy outcomes, trimesters, and complications (Supplemental Table 1). Gestational age (GA) was extracted from a prioritized code hierarchy and the start date was back-calculated from the encounter date minus the GA (Supplemental Material). Episode construction followed a dual-arm strategy. In the primary arm, encounters bearing a definitive outcome code served as anchors. In the second arm, remaining pregnancy encounters were grouped using gap-based heuristics. Each episode was then summarized with its prioritized outcome type and date, GA at outcome, estimated last menstrual period (LMP), and multiple-gestation type.

### 3.3. LLM prompting

Second, we developed an end-to-end LLM pipeline operating exclusively on unstructured notes and messages. For each patient, all available notes and messages were chronologically concatenated and provided as input to the model. The prompt instructed the LLM to detect each distinct pregnancy with its fetuses documented in the notes, to consolidate information across notes, to use dates and narrative context to temporally order pregnancies, and to output one structured JSON object per fetus containing start date, end date, gestational age, outcome, complications, delivery method, and assisted reproductive treatment when mentioned (Supplemental Material). The prompt asks for fetus-level information to allow for opposing outcomes within one pregnancy and individual fetal abnormality tracking. Pregnancy-level information was derived by taking the maximum across fetuses within each pregnancy (i.e. pregnancy end date was the date that the last fetus reached an outcome). The LLM prompt was engineered on training data to match human annotations. We used GPT-5 (version 2025-08-07; OpenAI, San Francisco, CA) with a training data cutoff of October 2024 with medium reasoning effort which was accessed via a Health Insurance Portability and Accountability Act (HIPAA)-compliant institutional cloud endpoint.

### 3.4. Manual chart review

Full manual chart-review with July 2025 as censor date served as the gold standard. Patients were selected based on structured pregnancy eras, with deliberate oversampling of difficult cases (low-confidence episodes, complications, loss of follow-up). The annotator reviewed the complete longitudinal record (clinical notes, laboratory results, procedure reports, and patient messages) following a written protocol that defined terms and rules (Supplemental Material). Annotated variables included pregnancy start date, end date, gestational age, outcome (live birth, spontaneous or induced abortion, stillbirth), complications (gestational diabetes, hyperemesis), and use of assistive reproductive technology (in vitro fertilization (IVF), intrauterine insemination (IUI)). We annotated individual fetuses rather than whole pregnancies, which allowed fetal abnormalities and opposing outcomes within one pregnancy to be tracked separately. Chart review was labor-intensive, requiring more than 30 minutes per patient because documentation was fragmented across many notes and encounters. Annotation was performed by a single annotator (S.A., clinical research coordinator) for an initial exploratory set (4 patients, 11 pregnancies) for protocol development, a training set (10 patients, 20 pregnancies) for prompt engineering and protocol adaptation, and a held-out test set (20 patients, 54 pregnancies) (Table 1). Pregnancy-level information was derived in the same way as for the LLM output.

**Table 1.** Cohort description.

| **Characteristic** | **Value** |
|---|---|
| Total patients (train/test) | 30 (10/20) |
| Number of pregnancy episodes (range per person) (train/test) | 74 (1 - 10) (20/54) |
| Number of fetuses (train/test) | 76 (21/55) |
| Study period | 1997-01-01 to 2025-04-20 |
| **Pregnancy outcomes, n (%)** |  |
| Live birth | 42 (56.8) |
| Stillbirth | 1 (1.4) |
| Spontaneous abortion | 14 (18.9) |
| Induced abortion | 10 (13.5) |
| Abortion NOS | 5 (6.8) |
| Ectopic pregnancy | 0 (0.0) |
| Molar pregnancy | 0 (0.0) |
| Unknown | 2 (2.7) |
| **Delivery method of live births, n (%)** |  |
| Spontaneous vaginal | 9 (21.4) |
| Induced vaginal | 12 (28.6) |
| Scheduled cesarean | 5 (11.9) |
| Emergency cesarean | 8 (19) |
| Unknown/Not recorded | 8 (19) |
| **Assisted reproductive treatment, n (%)** |  |
| IVF | 9 (12.2) |
| IUI | 0 (0.0) |
| Other/Unknown | 0 (0.0) |
| **Maternal complications, n (%)** |  |
| Gestational hypertension | 8 (10.8) |
| Preeclampsia | 3 (4.1) |
| Gestational diabetes | 5 (6.8) |
| Postpartum hemorrhage | 5 (6.8) |
| Placental abnormalities | 5 (6.8) |
| **Data completeness, n (%)** |  |
| Missing outcome date | 12 (16.2) |
| Missing estimated start date | 18 (24.3) |
| Pregnancy not followed at UCSF | 30 (40.5) |

The table lists characteristics of the manually annotated cohort. Percentages may not sum to 100 due to rounding.
NOS: not otherwise specified, IVF: in-vitro fertilization, IUI: intrauterine insemination

### 3.5. Comparison of approaches to the gold standard

LLM- and structured-data-derived pregnancy episodes were compared against chart-review at the pregnancy level, with fetus-level information aggregated per pregnancy; LLM and chart-review were additionally compared at the fetus level. Predicted and annotated pregnancy episodes of the same patient were matched one-to-one in a greedy procedure: annotated episodes with known dates were processed first, and each was assigned the unmatched predicted episode with the smallest absolute difference between any pair of start and end dates, provided that the start or end date differed by at most 365 days. Outcome type was used only as a fallback criterion for annotated episodes without a known outcome date. As a sensitivity check, using the sum of start- and end-date differences as the matching score yielded identical matches. Matched pregnancy episodes counted as true positives, unmatched annotated episodes as false negatives (missed), and unmatched predicted episodes as false positives (additional), from which we computed precision, recall, and F1 score. All comparisons between the two methods (dates, outcomes, complications, assisted reproductive treatment, multiple gestation) were restricted to pregnancies matched by both methods; pregnancies matched by only one method and unmatched predicted episodes were therefore not part of these paired analyses. Supplemental Table 7 lists the unit (patient, pregnancy, or fetus) and denominator of each analysis. We computed absolute error for start and outcome dates, and precision, recall, and F1 score for outcomes, complications, assisted reproductive treatment, and multiple gestation. Superiority of the LLM was tested with McNemar’s test for binary outcomes and the two-sided Wilcoxon signed-rank test for dates (p<0.05, uncorrected for multiplicity).

On the training set we conducted error analyses that informed prompt engineering and protocol adaptation. For this, we manually reviewed the LLM input of missed or unmatched pregnancies and examined misalignments in complication annotations by reviewing the evidence that the LLM provided and reexamining the complete chart.

## 4. Results

### 4.1. Cohort characteristics

Manual review of 30 patient records sampled from a pregnancy cohort identified from structured data revealed a total of 74 pregnancies (76 fetuses, including two twin pregnancies) over the study period (1997-2025; Table 1). In several cases, clinically relevant pregnancy information was present only in narrative notes or patient messages, with insufficient or absent structured coding due to care being carried out outside of UCSF. Historical pregnancies and assisted reproductive treatments were frequently documented in free text (e.g., GTPAL notation) but not captured in structured fields. For 12 pregnancies outcome dates could not be ascertained due to loss to follow-up and censoring. Start dates could not be determined for 18 pregnancies due to missing information on gestational age or outcome dates. The cohort includes 30 pregnancies not followed at UCSF but only mentioned in notes.

### 4.2. Training data results and prompt engineering

Prompt engineering of GPT-5 (Supplemental Material) and refinement of the ICD-code algorithm (Supplemental Table 1, Supplemental Figure 1) were performed on the training data by iteratively inspecting misalignments with manual annotation. The LLM produced no unmatched episodes (false positives) and the structured algorithm eight, the latter arising from repeated ICD code entries, isolated complication codes, or late postpartum documentation (pregnancy-level F1 0.97 vs 0.76; Supplemental Table 2). Of five LLM-manual disagreements on complications (Supplemental Table 3), two were human annotation errors, two stemmed from unspecific definitions, and one reflected information absent from the LLM input. The two definition issues were resolved by refining the protocol and prompt (one entry per fetus for multiple gestations; a definition of placental abnormality), after which four disagreements remained (two human errors, two due to incomplete LLM input). Further training-set results are in Supplemental Figures 2-4 and Supplemental Tables 3-5.

### 4.3. Validation of pregnancy episode detection

Of the 54 manually annotated test-set pregnancies, the LLM matched 53 and the structured algorithm 40 (Table 2; pregnancy-level F1 0.96 vs 0.68). The one pregnancy missed by the LLM was also missed by the structured algorithm, leaving 40 pregnancies matched by both methods, on which all further comparisons of the two methods are based; because records ended before the pregnancy did, the LLM labeled it ongoing rather than inferring an end date. The LLM produced 3 unmatched episodes (this pregnancy plus two extra pregnancies for a patient with 10 annotated pregnancies), whereas the structured algorithm produced 24, arising from duplicate ICD entries months apart, postpartum visits years after delivery, and isolated complication codes. The 13 test-set pregnancies matched only by the LLM are therefore not reflected in the date, outcome, and complication comparisons below.

**Table 2.** Matching pregnancies to manual chart review.

| **Method** | **True positive** | **False positive** | **False negative** | **Precision** | **Recall** | **F1** |
|---|---|---|---|---|---|---|
| LLM | 53 | 3 | 1 | 0.95 | 0.98 | 0.96 |
| Structured | 40 | 24 | 14 | 0.63 | 0.74 | 0.68 |

Note: The table shows per method the number of pregnancies that were successfully matched to human annotation (true positive), those that were missed (false negative), and additional ones (false positive) only identified by the LLM or structured data. A total of N=54 pregnancies were found in human annotation. Precision = TP/(TP+FP), recall = TP/(TP+FN), F1 = 2TP/(2TP+FP+FN). The number of predicted episodes (true plus false positives) differs between methods because each method generates its own candidate episodes; the reference set is identical for both. Abbreviations: FN, false negative; FP, false positive; LLM, large language model; TP, true positive.

### 4.4. Temporal accuracy of pregnancy episodes

LLM-derived start and end dates were closer to chart review than those derived from structured data (Figure 1). Date errors were compared for the 32 pregnancies with start and end dates available from all three sources. For the start date, the median LLM error was 0 days versus 25.5 for the structured data algorithm (two-sided Wilcoxon signed rank test: p-value < 0.001, W = 16). For the end date, the median LLM error was 0 days versus 1 day for the structured data algorithm (p-value = 0.044, W = 80). The largest LLM errors for start date were 22 days and for end date 115 days. The larger difference for the end date was due to both human and LLM approximating a date solely based on a year. Structured-data errors were more variable with more outliers (maximum end date difference = 220 days, maximum start date difference = 178 days). For the 14 abortion pregnancies in this set, LLM start dates were within 16 days of chart review (median absolute error 0 days), whereas the structured algorithm placed the start too early in 13 of 14 (median absolute error 59 days, maximum 106 days), likely reflecting its default gestational-age assumptions for early pregnancy losses.

**Figure 1:**
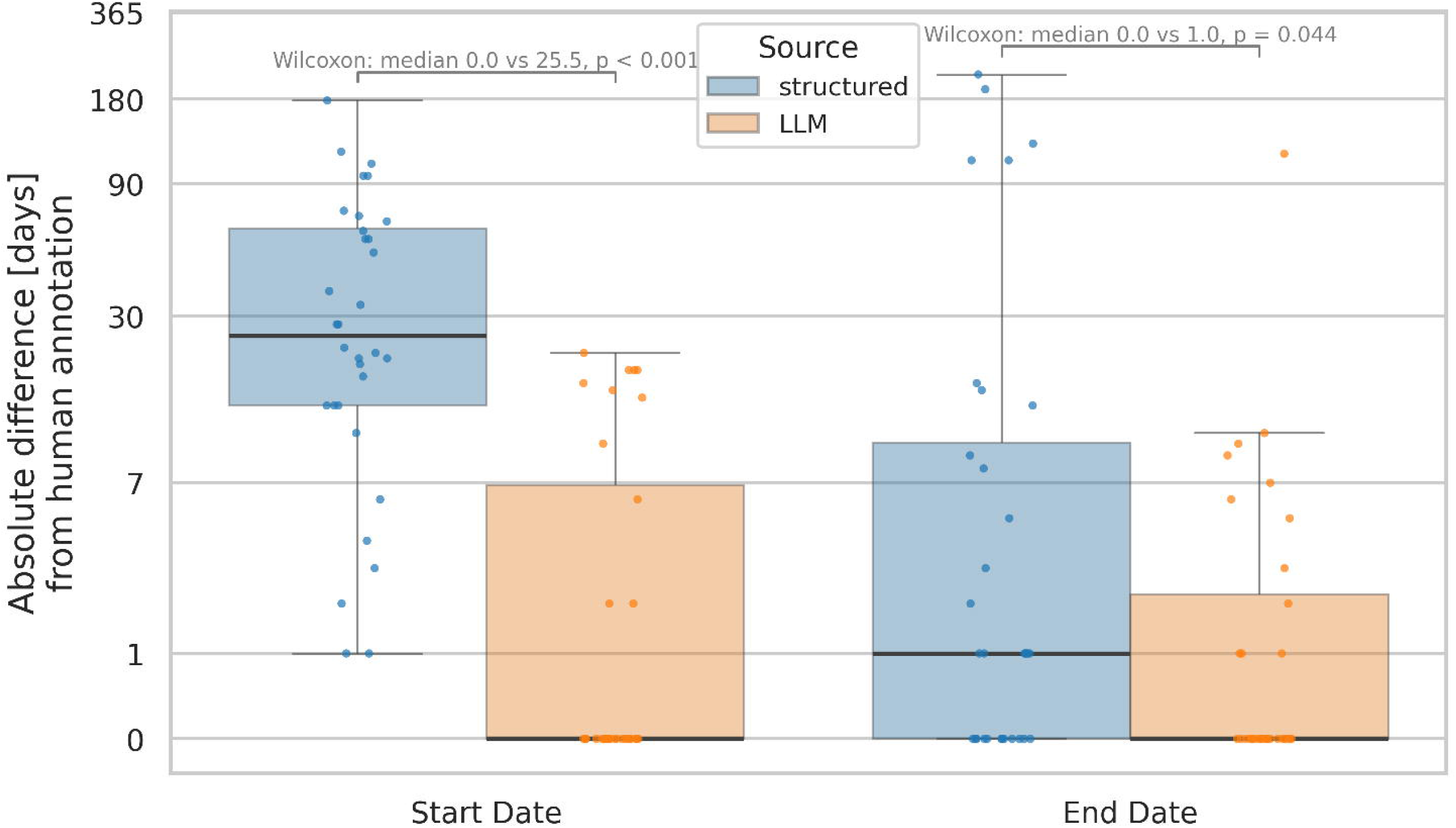
Temporal accuracy of pregnancies The boxplots depict for both start and end dates (x-axis) the absolute error in days (y-axis) for the structured algorithm and the LLM from human annotation. Shown are the 32 test-set pregnancies matched by both methods with start and end dates available from all sources. Individual dots display each pregnancy. The text annotation displays the results of the two-sided Wilcoxon signed rank test.

### 4.5. Pregnancy outcomes and complications

The LLM performed similarly to the structured-data algorithm in identifying pregnancy outcomes (McNemar’s test: p-value = 0.22, Figure 2, Supplemental Table 5). Of the 40 pregnancies matched by both methods, two had no annotated outcome because of the July 2025 censor date. Of the remaining 38, the LLM assigned the ground truth label for all but six pregnancies (accuracy = 0.84) while structured data misclassified 10 (accuracy = 0.74). Live births (N=19) were well identified by both methods (F1 score 0.94 for the LLM vs 0.91 for structured data) with some inaccuracies for “ongoing” in the LLM (N=2) and “unknown” for the structured algorithm (N=3).

**Figure 2:**
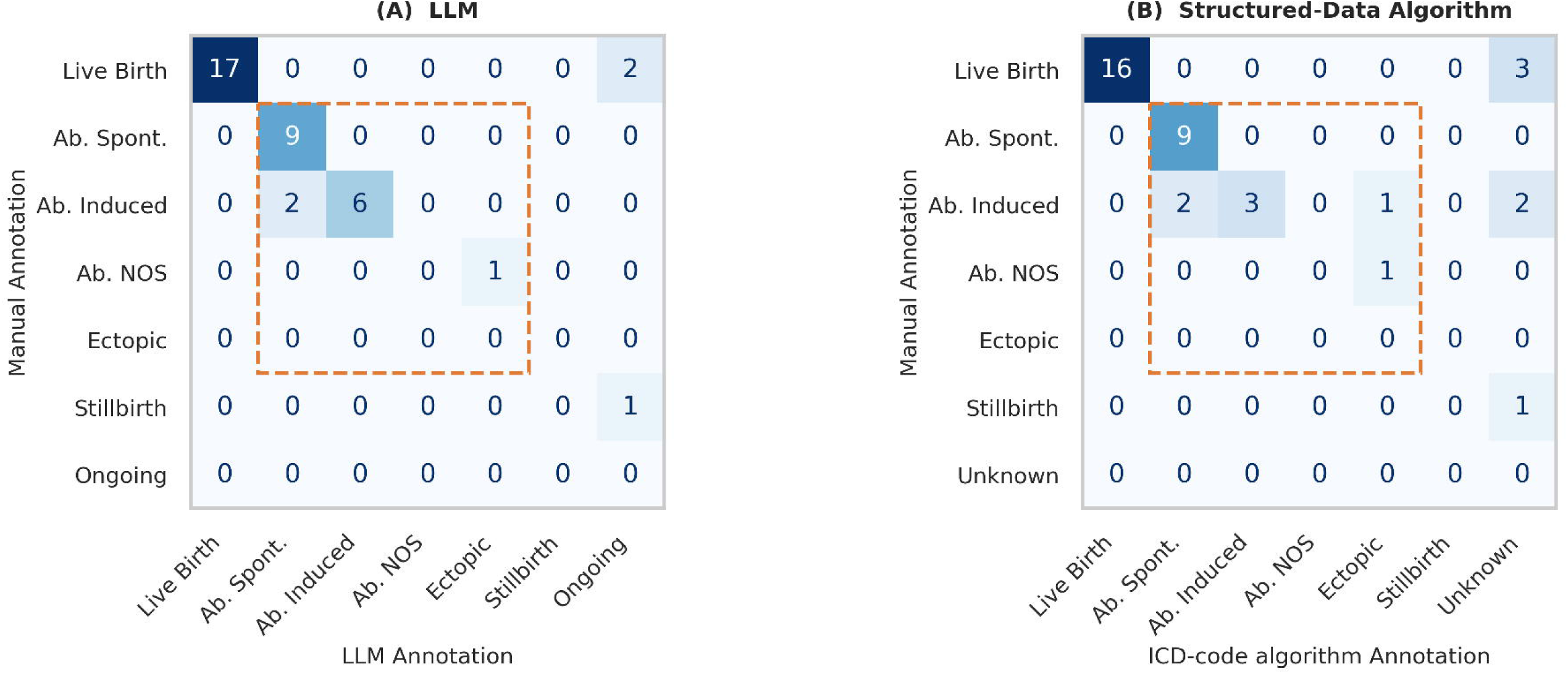
Performance on outcome types The performances of the two methods (LLM, structured-data algorithm) are shown for assigning the correct outcome type to a pregnancy episode. Shown are the 38 test-set pregnancies matched by both methods with an annotated outcome. The confusion matrices show the manual annotated outcome versus the predicted one, A) LLM, B) structured-data algorithm. The orange square indicates the abortion subcategories that were grouped together in the fuzzy matching. NOS: not otherwise specified, Ab.: Abortion

Specific abortion subtypes were more difficult to identify for both algorithms. Thus, we also calculated a “fuzzy” matching score, where induced, spontaneous, ectopic, and molar pregnancies formed a category (N=18; F1 score 1.00 for the LLM vs 0.94 for structured data). Both algorithms performed numerically better against this more lenient metric, with an accuracy on outcome classification of 0.84 for the structured algorithm and 0.92 for the LLM (McNemar’s test: p-value = 0.38, Supplemental Table 5).

Complications were frequently described in narrative text but inconsistently coded in structured data. Prevalence of the complications was low in our annotated dataset ranging from 0 (hyperemesis, placenta previa) to 0.1 (gestational diabetes), limiting statistical stability of per-condition estimates. Evaluated on the 40 pregnancies where both LLM and structured-data algorithm had matches (400 pregnancy-complication pairs), LLM achieved 0.8 recall and 0.84 precision pooled across all complications, compared with the structured-data algorithm’s 0.8 recall and 0.38 precision (McNemar’s test: p-value < 0.001, Figure 3, Supplemental Table 6). Fetal abnormalities showed the largest difference, because the structured algorithm counted any visit or consultation for a fetal abnormality, a choice made necessary by incomplete mother–fetus record linkage. Out of the 8 individual complications that were present in the dataset, the LLM achieved a higher or equal F1 score for 6. Only preeclampsia and chorioamnionitis were better identified by structured data, suggesting that a combination of both methods could improve performance for some complications.

**Figure 3:**
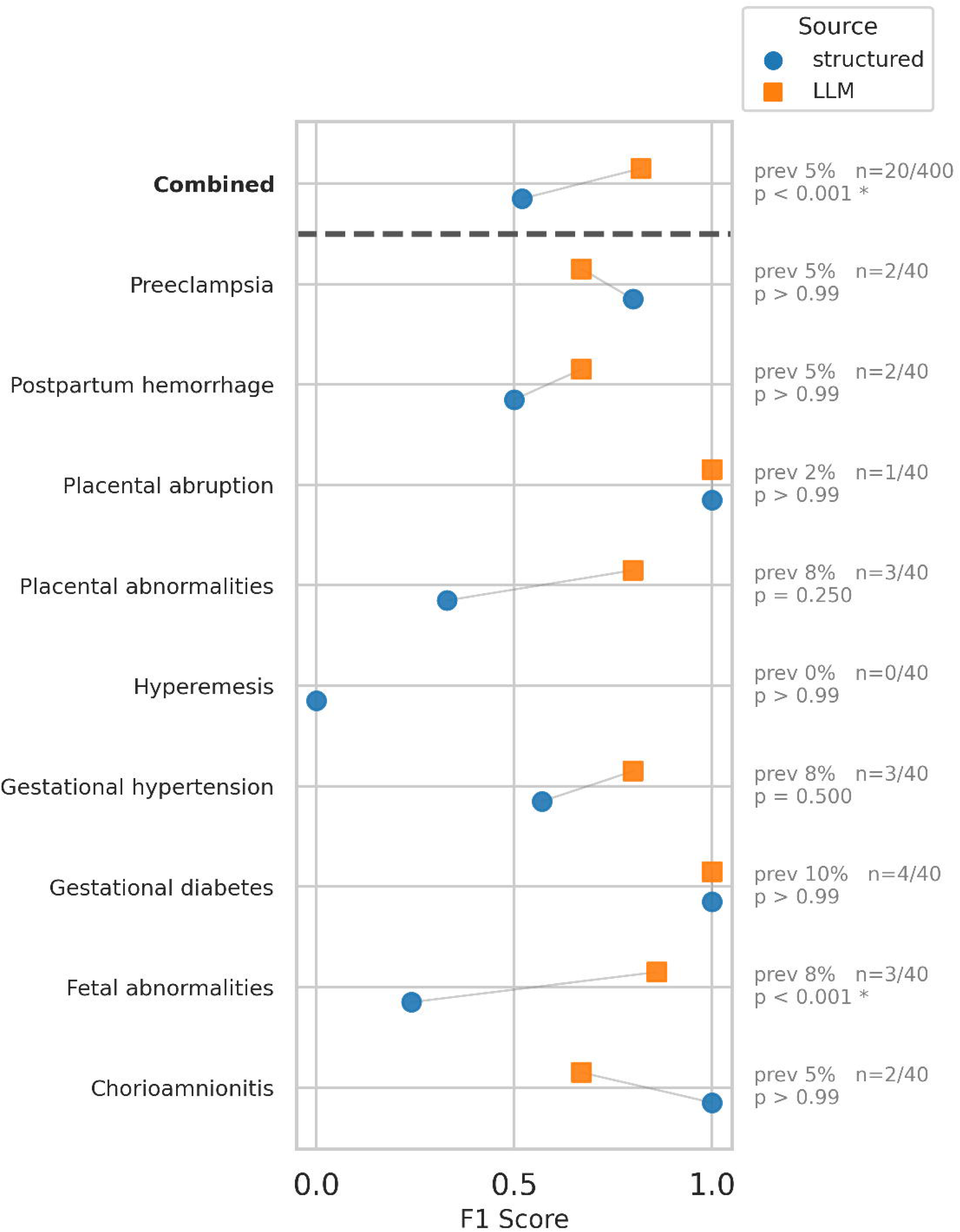
Performance of complication determination The barplot depicts the F1 score (x-axis) for each complication associated with the pregnancy (y-axis) and the combined performance across all complications. Evaluated on the 40 test-set pregnancies matched by both methods (combined: 400 pregnancy-complication pairs). The text on the right side denotes the prevalence of each complication as well as the respective McNemar’s test p-value. Complications where no true positives also no false positives were present, were removed from this figure (placenta previa). Hyperemesis had no positive cases; the structured algorithm’s F1 score of 0 reflects one false positive. Prev: prevalence

### 4.6. Information unavailable in structured data

ICD code O09.81 records that assisted reproduction was used but not which treatment, whereas notes documented the treatment type explicitly and the LLM extracted it. On the test set, the LLM achieved 100% accuracy for assisted reproductive treatment identification (among the 40 pregnancies matched by both methods: 7 true positives, 33 true negatives, one of which the LLM labelled “unknown”; among all 53 LLM-matched pregnancies: 7 true positives, 46 true negatives). In our dataset, only IVF was present. Given the small number of positive cases, this estimate is imprecise. The binary identification of whether the pregnancy was a result of any assistive reproductive treatment was possible from structured data and achieved 100% accuracy (N=40 pregnancies matched by both methods; 7 true positives, 33 true negatives).

The LLM-based approach also allows fetus-level tracking. The test data included one multiple gestation as did the train data. Identification of whether a pregnancy was a multiple gestation was possible from structured data though one false positive was identified in the test set (N=40; 1 true positive, 1 false positive, 0 false negatives; accuracy = 0.975, precision = 0.50, recall = 1.00). The LLM correctly classified all pregnancies, including the single multiple gestation, and identified its number of fetuses; with only one multiple gestation in the test set, this result should be interpreted with caution. While rare, in the train data the twin pregnancy did have different outcomes for the two fetuses with one fetal demise and one live birth. The LLM identified these opposing outcomes and assigned the correct outcome date for the live birth fetus and a slightly earlier date to the fetus that died in utero (difference to manual annotation: 12 days).

## 5. Discussion

When benchmarked against manual chart-review, the LLM-based approach identified more pregnancy episodes with fewer spurious episodes, estimated start and end dates more accurately, and identified complications with higher precision than the structured-data pregnancy algorithm; outcome classification was numerically but not significantly better. Importantly, the LLM was able to extract important clinical information unavailable in structured EHR data, highlighting the added value of clinical notes for pregnancy episode phenotyping in real-world data settings.

Prior pregnancy identification methods relying on diagnosis and procedure codes have reported high positive predictive value for live births and delivery dates, particularly in administrative claims and well-curated EHR datasets. However, they are known to struggle with accurate pregnancy start-date estimation, early pregnancy losses, non-live-birth outcomes^1,3–9^, and pregnancies not fully followed within a single health system. Our own implementation of a hierarchical algorithm based on structured data reproduced many of these known behaviors, including the creation of spurious or fragmented episodes driven by repeated postpartum codes, isolated complication codes, or delayed coding practices and inaccuracies in start dates.

The presented LLM-based approach addresses these data limitations by inferring pregnancy episodes from clinician narratives rather than from coded events. As a result, the notes-based pipeline was more robust to fragmented care patterns and more closely matched full chart-review. This is consistent with prior observations that structured data approaches trade generalizability and completeness for simplicity and scalability, and that incorporating clinical narrative improves performance^19^. Importantly, our comparison does not separate the contribution of the input data from that of the model: the LLM received longitudinal notes and messages, whereas the comparator was restricted to ICD codes, and 30 of the 74 annotated pregnancies were documented only in narrative text. The observed gains therefore reflect the combined LLM-plus-narrative pipeline rather than LLM reasoning alone. Best practice for computable phenotypes combines structured and narrative data, and our complication results (e.g., preeclampsia, chorioamnionitis) suggest that such hybrid approaches may perform best.

LLMs can also make previously inaccessible information available. The type of assisted reproductive treatment cannot be determined from ICD codes, and opposing outcomes within a multiple gestation are hard to extract when mother-infant records are not linked, yet both are documented in clinical narratives. Compared with traditional NLP, LLMs need less annotated data and can extract more complex variables, which could be written back to structured databases for research and care, for example to fill gaps in the records of patients new to a health system.

Despite improved performance, LLM-based extraction remains fundamentally constrained by the completeness of available documentation. Most missed or unmatched pregnancies were attributable to missing or inaccessible documentation rather than model failure, although one test-set pregnancy was missed because the LLM treated a pregnancy with incomplete follow-up as ongoing. This emphasizes the importance of improving data capture and sharing^20^.

The training-phase error analysis revealed that human annotation errors occurred at a comparable frequency to LLM errors, highlighting that pregnancy phenotyping itself is inherently ambiguous and difficult like several other clinical annotation tasks^21^. This underscores the importance of clear annotation protocols and alignment between human reviewers and model prompt, analogous to inter-annotator agreement considerations in traditional NLP evaluations^21^.

LLM-based approaches also pose distinct reproducibility challenges: outputs depend on prompt design, model version, and settings such as reasoning effort or temperature. Whereas rule-based algorithms are deterministic once specified, LLM pipelines require prompt versioning, output validation, and monitoring for distribution shift^22^, as well as consistency checks^23^.

Published rule-based pregnancy algorithms are often difficult to transfer across institutions due to differences in coding practices, data models, and care workflows. While LLMs lower the barrier to adaptation by relying on natural language rather than local code sets, they are not inherently plug-and-play, because note structure, abbreviations, and documentation culture differ between sites. Nevertheless, adapting an LLM prompt may be less labor-intensive than re-engineering a large rule-based phenotype, but whether this translates into better portability remains to be shown.

This study has several limitations. First, the sample size of the annotated data was modest, reflecting the intensive nature of full manual chart-review, and limits the robustness of performance estimates. Patients were also deliberately enriched for difficult cases (low-confidence episodes, complications, loss to follow-up), so our estimates should not be read as performance in an unselected obstetric population, and results for rare features (assisted reproductive treatment, multiple gestation, individual complications) rest on very few cases. Second, manual review was performed by a single annotator, precluding assessment of inter-annotator agreement. Third, data were drawn from a single academic medical center, and documentation practices at UCSF may not reflect those of other health systems. The cohort may also be biased toward more complicated pregnancies, because it consisted of patients with inflammatory bowel disease receiving tertiary care, whose documentation and pregnancy risks may differ from the general population. Fourth, patients contributed multiple pregnancies and complications were evaluated jointly within pregnancies, whereas our statistical tests treated observations as independent; p-values should therefore be interpreted with caution. Fifth, for annotated episodes without a known outcome date, outcome type served as a fallback matching criterion, which may slightly favor outcome agreement for these episodes (4 of 53 LLM-matched and none of 40 structured-matched test-set pregnancies). Sixth, paired comparisons were restricted to pregnancies matched by both methods, which excludes the pregnancies found only by the LLM and may favor the structured algorithm.

## 6. Conclusions

LLM-based pregnancy episode extraction is a promising complement to code-based pregnancy phenotyping in real-world data. Future work should scale evaluation to larger, more diverse populations, incorporate multiple annotators, and assess robustness across institutions and EHR systems. Hybrid approaches that combine structured signals with LLM-based note interpretation should be developed and evaluated, as they are likely to perform best. More broadly, LLMs applied to longitudinal clinical narratives can support inference beyond entity extraction, enabling richer use of unstructured EHR data for reproductive health research.

### Clinical Relevance Statement

Pregnancy cohorts built from diagnosis codes alone can miss pregnancies, misplace start dates by weeks, and lose details such as assisted reproduction or fetus-level outcomes, which can distort studies of exposures during pregnancy. In this proof-of-concept study, an LLM applied to longitudinal clinical notes identified pregnancies and their dates in close agreement with manual chart review. Teams building pregnancy cohorts from electronic health record data should consider pairing code-based algorithms with LLM review of clinical notes, after validating performance locally.

### Protection of Human and Animal Subjects

The study was performed in compliance with the World Medical Association Declaration of Helsinki on Ethical Principles for Medical Research Involving Human Subjects and the U.S. Common Rule (45 CFR 46), and was reviewed and approved by the University of California, San Francisco Institutional Review Board (Study #24-43008).

The UCSF Institutional Review Board granted a waiver of informed consent for this retrospective study, as it posed no more than minimal risk and could not practicably be conducted without the waiver or without access to identifiable information.

## Author contributions

A.S.: Conceptualization, Data analysis, Writing - Original Draft. S.A.: Manual Annotation, Error analysis, Writing - Review & Editing. A.A: Writing - Review and Editing, A.B.: Writing - Review and Editing. M.W.: Conceptualization, Writing - Review and Editing. V.R.: Conceptualization, Supervision, Writing - Review & Editing.

## Acknowledgments

This publication is supported by the Food and Drug Administration (FDA) of the U.S. Department of Health and Human Services (HHS) as part of a financial assistance award Center of Excellence in Regulatory Science and Innovation grant to University of California, San Francisco, U01FD005978 totaling $ 350,000. The contents are those of the author(s) and do not necessarily represent the official views of, nor an endorsement, by FDA/HHS, or the U.S. Government. The authors thank the UCSF AI Tiger Team, Academic Research Services, Research Information Technology, and the Chancellor’s Task Force for Generative AI for their software development, analytical, and technical support related to the use of Versa API gateway (the UCSF secure implementation of large language models and generative AI via API gateway). We further thank the Facility for Advanced Computing (FAC) at UCSF for access to compute and data storage resources.

## Data Availability

The data is protected health information and will thus not be made publicly available. The LLM prompt is provided in the Supplemental Material.

## Code Availability

The analysis code is not publicly available.

## Disclaimer

This article reflects the best judgement of the authors and should not be construed to represent FDA’s views or policy.

## Multiple-Choice Questions

Question 1. Which pregnancy attribute showed the largest accuracy gap between the ICD-code algorithm and the LLM applied to clinical notes?

A. Identification of live births
B. Pregnancy start date
C. Pregnancy end date
D. Multiple gestation status

## Correct answer

B. Pregnancy start date

## Explanation

Among the 32 test-set pregnancies with dates available from all sources, the median absolute error for the start date was 0 days for the LLM and 25.5 days for the ICD-code algorithm (p < 0.001). Code-based algorithms have to back-calculate the start of a pregnancy from gestational-age codes or default assumptions, which is especially error-prone for early pregnancy losses: for the 14 abortion pregnancies, the structured algorithm placed the start too early in 13, with a median error of 59 days.

By contrast, end dates differed by a median of only 1 day, live births were identified well by both methods, and the test set contained only one multiple gestation, so none of these showed a comparable gap.

Question 2. Which pregnancies are most likely to be missed by algorithms that rely only on diagnosis and procedure codes?

A. Pregnancies delivered at the index hospital
B. Pregnancies cared for at outside institutions
C. Pregnancies complicated by gestational diabetes
D. Pregnancies ending in cesarean delivery

## Correct answer

B. Pregnancies cared for at outside institutions

## Explanation

Diagnosis and procedure codes are generated when care is delivered and billed within the health system, so pregnancies managed elsewhere, or mentioned only as obstetric history, leave little or no coded trace. In this cohort, 30 of the 74 annotated pregnancies were not followed at the study institution and were documented only in narrative notes, often in obstetric shorthand such as GTPAL.

Deliveries at the index hospital and gestational diabetes generate codes during in-system care and are captured reasonably well by structured data.

Question 3. What is the main reason the performance gains in this study reflect both the data source and the language model?

A. The prompt was refined on the held-out test set
B. The two methods were given different input data
C. Chart review was performed by several annotators
D. The LLM also received structured diagnosis codes

## Correct answer

The two methods were given different input data

## Explanation

The LLM received longitudinal clinical notes and patient messages, whereas the comparator algorithm used only ICD codes, and 30 of the 74 annotated pregnancies were documented only in narrative text. The comparison therefore measures the combined effect of a richer data source and the model, rather than the model alone.

The other options are incorrect: the prompt was engineered on a separate training set, chart review was performed by a single annotator, and the LLM pipeline used only notes and messages.

## References

1 Blotière P, Weill A, Dalichampt M, et al. Development of an algorithm to identify pregnancy episodes and related outcomes in health care claims databases: An application to antiepileptic drug use in 4.9 million pregnant women in France. Pharmacoepidemiol Drug Saf 2018;27:763–770. doi:10.1002/pds.4556

2 Smith LH, Wang W, Keefe-Oates B. Pregnancy episodes in All of Us: harnessing multi-source data for pregnancy-related research. J Am Med Inform Assoc 2024;31:2789–2799. doi:10.1093/jamia/ocae195

3 Canelón SP, Burris HH, Levine LD, et al. Development and evaluation of MADDIE: Method to Acquire Delivery Date Information from Electronic health records. Int J Med Inform 2021;145:104339. doi:10.1016/j.ijmedinf.2020.104339

4 Hornbrook MC, Whitlock EP, Berg CJ, et al. Development of an Algorithm to Identify Pregnancy Episodes in an Integrated Health Care Delivery System. Health Serv Res 2007;42:908–927. doi:10.1111/j.1475-6773.2006.00635.x

5 Ailes EC, Zhu W, Clark EA, et al. Identification of pregnancies and their outcomes in healthcare claims data, 2008–2019: An algorithm. PLoS One 2023;18:e0284893. doi:10.1371/journal.pone.0284893

6 Jones SE, Bradwell KR, Chan LE, et al. Who is pregnant? Defining real-world data-based pregnancy episodes in the National COVID Cohort Collaborative (N3C). JAMIA Open 2023;6:ooad067. doi:10.1093/jamiaopen/ooad067

7 Lyu T, Liang C, Liu J, et al. Temporal Events Detector for Pregnancy Care (TED-PC): A rule-based algorithm to infer gestational age and delivery date from electronic health records of pregnant women with and without COVID-19. PLoS One 2022;17:e0276923. doi:10.1371/journal.pone.0276923

8 Naleway AL, Gold R, Kurosky S, et al. Identifying pregnancy episodes, outcomes, and mother-infant pairs in the Vaccine Safety Datalink. Vaccine 2013;31:2898–2903. doi:10.1016/j.vaccine.2013.03.069

9 Matcho A, Ryan P, Fife D, et al. Inferring pregnancy episodes and outcomes within a network of observational databases. PLoS One 2018;13:e0192033. doi:10.1371/journal.pone.0192033

10 Zhong Q-Y, Mittal LP, Nathan MD, et al. Use of natural language processing in electronic medical records to identify pregnant women with suicidal behavior: towards a solution to the complex classification problem. Eur J Epidemiol 2019;34:153–162. doi:10.1007/s10654-018-0470-0

11 Belsti Y, Moran L, Mousa A, et al. Analyzing electronic medical records to extract prepregnancy morbidities and pregnancy complications: Toward a learning health system. Learn Health Syst 2024;9:e10473. doi:10.1002/lrh2.10473

12 Alsentzer E, Rasmussen MJ, Fontoura R, et al. Zero-shot interpretable phenotyping of postpartum hemorrhage using large language models. NPJ Digit Med 2023;6:212. doi:10.1038/s41746-023-00957-x

13 Huang J, Yang DM, Rong R, et al. A critical assessment of using ChatGPT for extracting structured data from clinical notes. NPJ Digit Med 2024;7:106. doi:10.1038/s41746-024-01079-8

14 Patel PV, Davis C, Ralbovsky A, et al. Large Language Models Outperform Traditional Natural Language Processing Methods in Extracting Patient-Reported Outcomes in Inflammatory Bowel Disease. Gastro Hep Adv 2025;4:100563. doi:10.1016/j.gastha.2024.10.003

15 Yim R, Silverman A, Wang S, et al. Optimising large language models for clinical information extraction: a benchmarking study in the context of ulcerative colitis research. BMJ Digit Health AI 2025;1:e000014. doi:10.1136/bmjdhai-2025-000014

16 He J, Rasmy L, Li H, et al. Prompting large language models for clinical temporal relation extraction. arXiv [preprint]. 2024. doi:10.48550/arXiv.2412.04512

17 Wang J, Weiss JC. A Large-Language Model Framework for Relative Timeline Extraction from PubMed Case Reports. AMIA Jt Summits Transl Sci Proc 2025;2025:598–606

18 Kougia V, Sedova A, Stephan AJ, et al. Analysing zero-shot temporal relation extraction on clinical notes using temporal consistency. In: Proceedings of the 23rd Workshop on Biomedical Natural Language Processing. Bangkok, Thailand: Association for Computational Linguistics; 2024:72–84

19 Moldwin A, Demner-Fushman D, Goodwin TR. Empirical Findings on the Role of Structured Data, Unstructured Data, and their Combination for Automatic Clinical Phenotyping. AMIA Jt Summits Transl Sci Proc 2021;2021:445–454

20 Tayefi M, Ngo P, Chomutare T, et al. Challenges and opportunities beyond structured data in analysis of electronic health records. Wiley Interdiscip Rev Comput Stat 2021;13:e1549. doi:10.1002/wics.1549

21 Chapman WW, Dowling JN, Hripcsak G. Evaluation of training with an annotation schema for manual annotation of clinical conditions from emergency department reports. Int J Med Inform 2008;77:107–113. doi:10.1016/j.ijmedinf.2007.01.002

22 Bommasani R, Hudson DA, Adeli E, et al. On the opportunities and risks of foundation models. arXiv [preprint]. 2021j. doi:10.48550/arXiv.2108.07258

23 Amodei D, Olah C, Steinhardt J, et al. Concrete problems in AI safety. arXiv [preprint]. 2016. doi:10.48550/arXiv.1606.06565

