## Supplemental materials for "Beyond billing codes: a proof-of-concept study of large language models for pregnancy episode reconstruction from clinical notes"

Supplemental Material

### ICD-code algorithm details

We implemented a hierarchical, rule-based algorithm (Supplemental Figure 1) to identify and characterize discrete pregnancy episodes from longitudinal ICD codes (Chapter O and Z32–Z37 ranges). ICD codes were mapped to pregnancy outcomes, trimesters, and complications (Supplemental Table 1). Gestational age (GA) was extracted from a prioritized code hierarchy: exact Z3A week-of-gestation codes, followed by trimester-specific suffix coding and named trimester indicator codes, and finally outcome-type midpoints (e.g., 36 weeks for preterm delivery codes). The start date was back-calculated from the encounter date minus GA-derived days; for encounters lacking GA, term (40 weeks), or early-encounter (10 weeks) assumptions were applied as fallbacks. Episode construction followed a dual-arm strategy. In the primary arm, encounters bearing a definitive outcome code served as anchors; concurrent outcome codes within a type-specific time window (28–120 days) were grouped, and a retrospective lookback window (GA-adjusted or outcome-type default, up to 300 days) was applied to capture all preceding prenatal encounters. Boundaries were refined by checking GA monotonicity, clinical consistency with the outcome type, and temporal continuity (encounters separated by >180 days were treated as separate episodes). In the second arm, remaining pregnancy encounters were grouped using gap-based heuristics (≤60 days: same episode; 60–180 days: clinical continuity check on GA progression and early-pregnancy markers; >180 days: new episode). Each episode was then summarized with hierarchy-defined prioritized outcome type, outcome date, GA at outcome, estimated LMP, episode duration, encounter frequency, and multiple-gestation type. This approach reflects commonly used structured EHR phenotyping strategies.

### LLM prompt details

#### Regex patterns to identify relevant MyChart and Staff messages

preg_terms = [

r'\bpregnan(t|cy)\b',

r'\bgravid\b',

r'\bgestation(al)?\b',

r'\bprenatal\b',

r'\bantepartum\b',

r'\bpost\s*partum\b',

r'\bvaginal\s+delivery\b',

r'\bc-section\b',

r'\bmiscarriage\b',

r'\bfetal\b',

r'\bfetus\b',

r'\btrimester\b',

r'\babortion\b',

r'\b(?:child ?birth|birth(?! ?control|date| date)|birth ?date|date of birth|DOB)\b',

r'\bgave birth\b',

r'\bchildbirth\b',

r'\bLMP\b',

r'\bdelivered\s+(the\sbaby|baby)\b',

r'\blast\s+menstrual\s+period\b',

r'\bdue\s+date\b',

r"\b(I'm|I am)\s+due\s+in\b",

r'\bD\s*&\s*C\b',

r'\bD\s*&\s*E\b',

]

#### Department specialty filter to get OBGYN notes

preg_provider_keywords = r'(?i)obstetrics|gynecology|prenatal|reproductive|maternal|fertility|fetal|women\'s health'

structured_flag = (

notes['department_specialty'].str.contains(preg_provider_keywords, na=False) |

notes['provider_specialty'].str.contains(preg_provider_keywords, na=False)

)

#### Prompt for GPT-5

You are an expert medical data extraction system specialized in analyzing obstetrics and gynecology notes.

Your task is to extract comprehensive information about each fetus and pregnancy mentioned in the clinical notes.

For each individual fetus of a pregnancy, extract the following:

1. Pregnancy ID: Assign a sequential number to each pregnancy (1 for earliest, 2 for next, etc.)

2. Fetus ID: Assign a sequential number to each fetus within a pregnancy (Pregnancy 1 might have Fetus 1 and Fetus 2 for twins)

3. Estimated start date: PRIORITIZE LMP when available. If multiple sources exist, use the following priority order: LMP > early ultrasound dating > stated gestational age with date > conception date > first detection.

4. First visit date: When pregnancy was first documented/first visit regarding this pregnancy

5. Estimated end date: When pregnancy ended (delivery date, miscarriage date, or expected end date)

6. Last visit date: When was pregnancy last discussed

7. Due date: If explicitly stated, extract it. If not stated, compute as 40 weeks from the start date.

8. Gestational age at end (weeks): Extract if explicitly stated. Otherwise compute from start and end dates when possible. If insufficient information, leave null.

9. Outcome: One of [live birth, stillbirth, spontaneous abortion/miscarriage, elective/induced abortion, abortion not otherwise specified, ectopic pregnancy, molar pregnancy, ongoing]

10. Delivery method (if applicable): [spontaneous vaginal, induced vaginal, planned cesarean, emergency cesarean, cesarean not otherwise specified, no delivery, unknown]

11. Fertility treatment: one of [None, IVF, IUI, Other, Unknown]

12. Maternal complications: List all maternal complications associated with the pregnancy, including but not limited to:

- Gestational diabetes

- Gestational hypertension

- Preeclampsia/eclampsia

- Hyperemesis gravidarum

- Placental abruption

- Placenta previa

- Chorioamnionitis

- Placental issues

- Premature rupture of membranes

- Preterm labor

- Intrauterine growth restriction

- Infections

- Postpartum hemorrhage

- Postpartum depression

Additionally, capture other complications not listed above under "other_complications".

13. Fetus-specific information:

- Sex (if mentioned)

- Birth weight (if mentioned)

- APGAR scores (if mentioned)

- Fetal/newborn complications

IMPORTANT INSTRUCTIONS:

- The patient notes may mention multiple pregnancies and potentially each with multiple fetuses. Create a separate entry for EACH DISTINCT fetus.

- A summary of all pregnancies is often given as GTPAL (gravidity, term, preterm, abortion, living). Use this as orientation. If GTPAL values differ, favor the version most frequently documented or most consistent with the narrative pregnancy history. Note that a twin pregnancy will appear twice under "TPAL".

- For multiple gestations (twins, triplets, etc.), create separate entries for each fetus with the SAME pregnancy_id but different fetus_id values (1, 2, 3...). Most fields are pregnancy-level data (identical for all fetuses), however, the following fields CAN differ between fetuses (end_date, gestational_age_weeks, outcome, sex, birth_weight, apgar_scores, fetal_complications).

- Combine all information that relates to the same fetus/pregnancy, even if it appears in different parts of the notes.

- Use the dates mentioned to organize pregnancies chronologically.

- Include direct quotes from the notes when relevant to support your extraction.

- If exact dates are not available, estimate based on contextual information (e.g., "last summer", "2 years ago"). Put 1900-01-01 if no date can be ascertained at all.

- Clearly distinguish between confirmed facts and likely inferences.

- For pregnancies with incomplete information, extract whatever details are available.

Output the information in the following JSON format:

{

"fetuses": [

{

"pregnancy_id": int,

"fetus_id": int,

"start_date": "string or null",

"first_visit_date": "string or null",

"end_date": "string or null",

"last_visit_date": "string or null",

"due_date": "string or null",

"gestational_age_weeks": float or null,

"outcome": "string or null",

"delivery_method": "string or null",

"fertility":"string or null",

"gestational_diabetes":int or null,

"gestational_hypertension":int or null,

"preeclampsia":int or null,

"hyperemesis_gravidarum":int or null,

"placental_abruption":int or null,

"placenta_previa":int or null,

"chorioamnionitis": int or null,

"placental_abnormalities":int or null,

"premature_rupture":int or null,

"preterm_labor":int or null,

"infection":int or null,

"postpartum_hemorrhage":int or null,

"postpartum_depression":int or null,

"fetal_abnormalities":int or null,

"other_complications": [

{

"complication_type": "string",

"details": "string",

"onset_date": "string or null"

}

],

"fetal_info": {

"sex": "string or null",

"birth_weight": "string or null",

"apgar_scores": "string or null",

"complications": "string or null"

},

"evidence": [

{

"text": "string",

"date_mentioned": "string or null"

}

]

}

]

}

"""

### Figures

#### Supplemental Figure 1: Flowchart of rules for the rule-based pregnancy episode algorithm

Visualization of how the pulled structured EHR data (ICD codes) were processed into pregnancy episodes. This included mapping of codes to gestational age, outcome type, and estimating the start date for each code if possible. Then constructing the episodes with a two-arm approach where definite outcomes serve as anchors and date thresholds are used to assign other ICD codes to them.

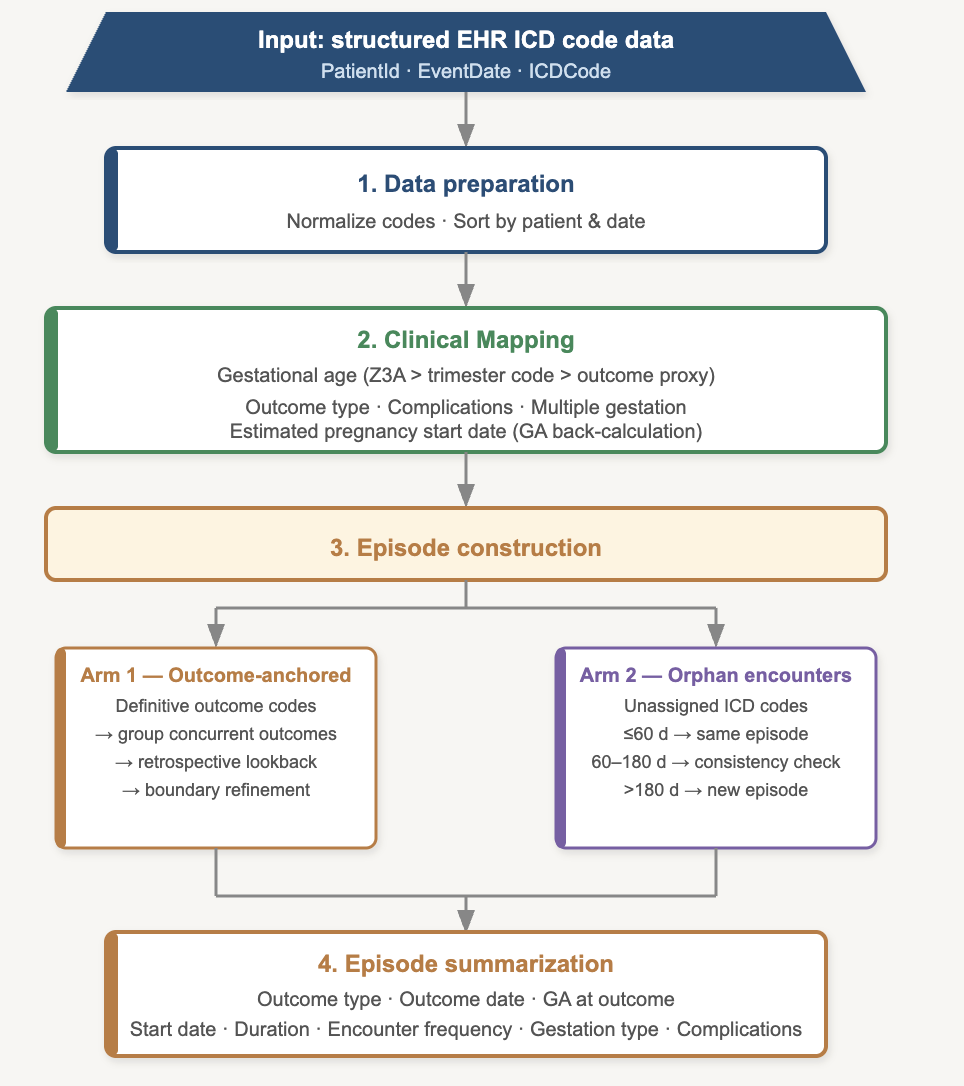

#### Supplemental Figure 2: Evaluation of start and outcome dates in training data

The boxplots depict for both start and outcome dates (x-axis) the absolute error in days (y-axis) for the structured algorithm and the LLM from human annotation. Individual dots display each pregnancy. The text displays each groups median and the two-sided Wilcoxon signed rank test. Shown are the 14 training-set pregnancies matched by both methods with start and end dates available from all sources.

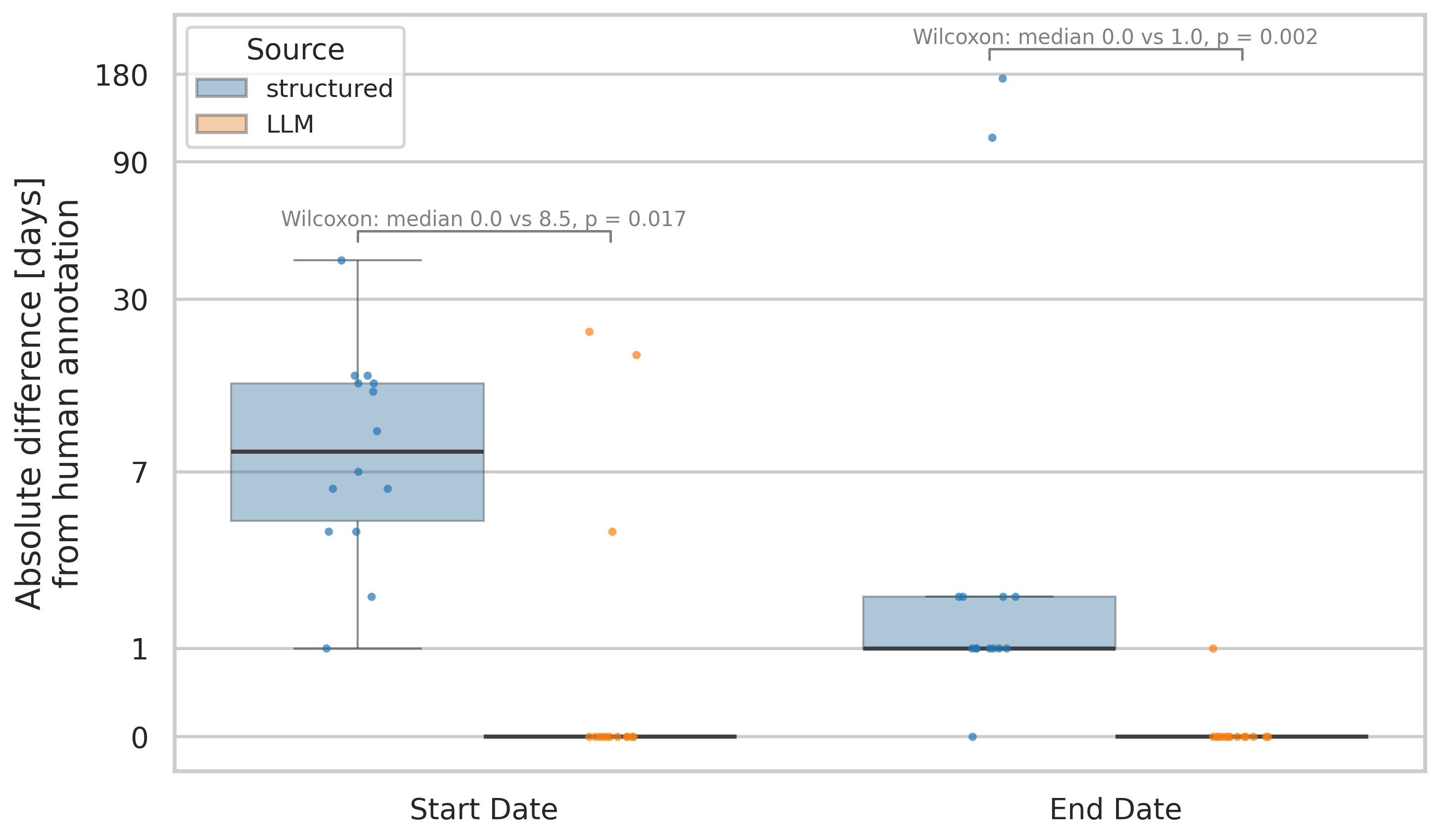

#### Supplemental Figure 3: Evaluation of complications on training data

The plot depicts the F1 score (x-axis) for each complication associated with the pregnancy (y-axis) and the combined performance across all complications. The text on the right side denotes the prevalence of each complication and the McNemar’s test p-value. Evaluated on the 17 training-set pregnancies matched by both methods (combined: 170 pregnancy-complication pairs).

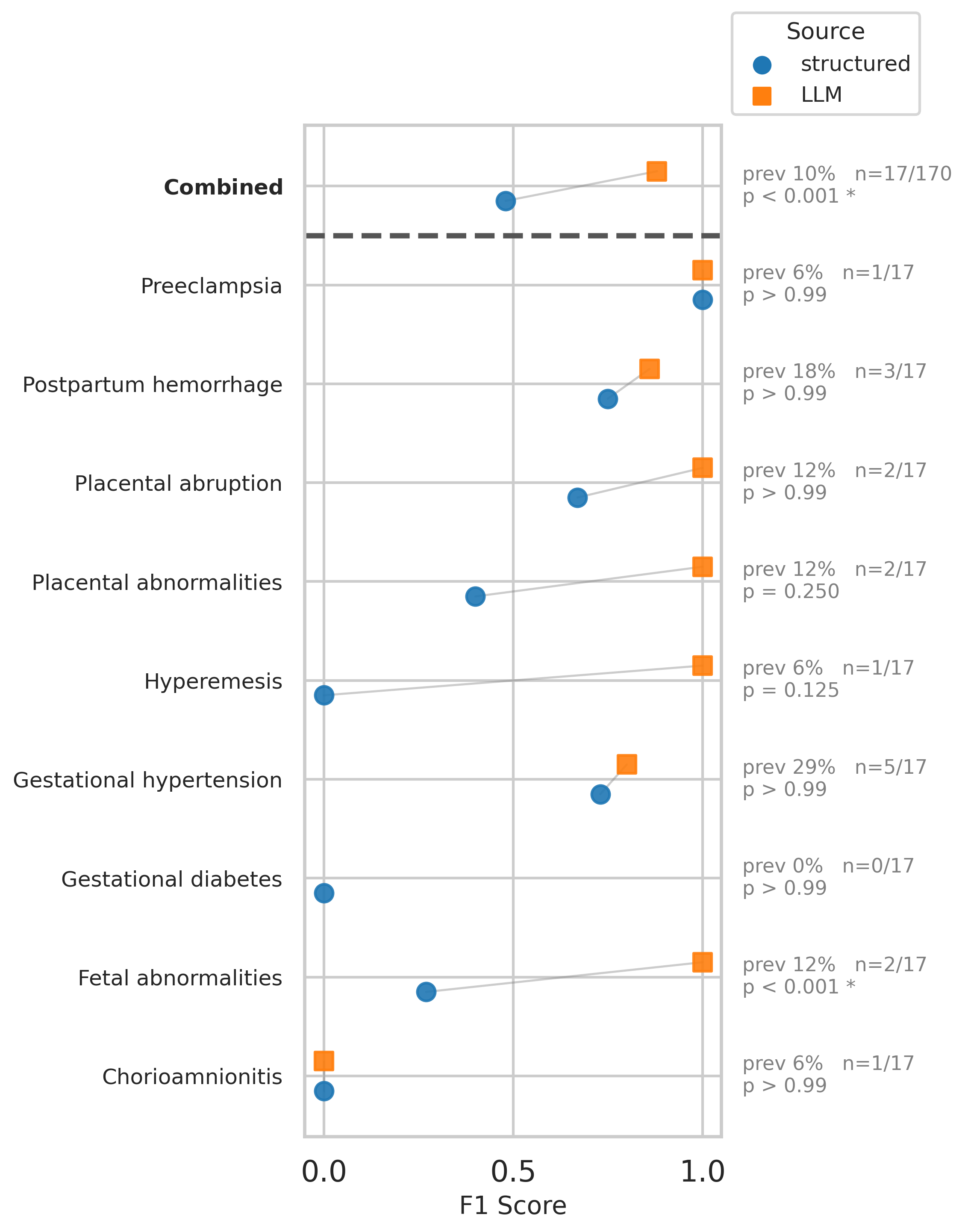

#### Supplemental Figure 4: Evaluation of outcome types on training data

The performance of the two methods (LLM, structured-data algorithm) are shown for assigning the correct outcome type to a pregnancy episode. The confusion matrices show the manual annotated outcome versus the predicted one, A) LLM, B) structured-date algorithm. The orange square indicates the abortion subcategories that were grouped together in the fuzzy matching.

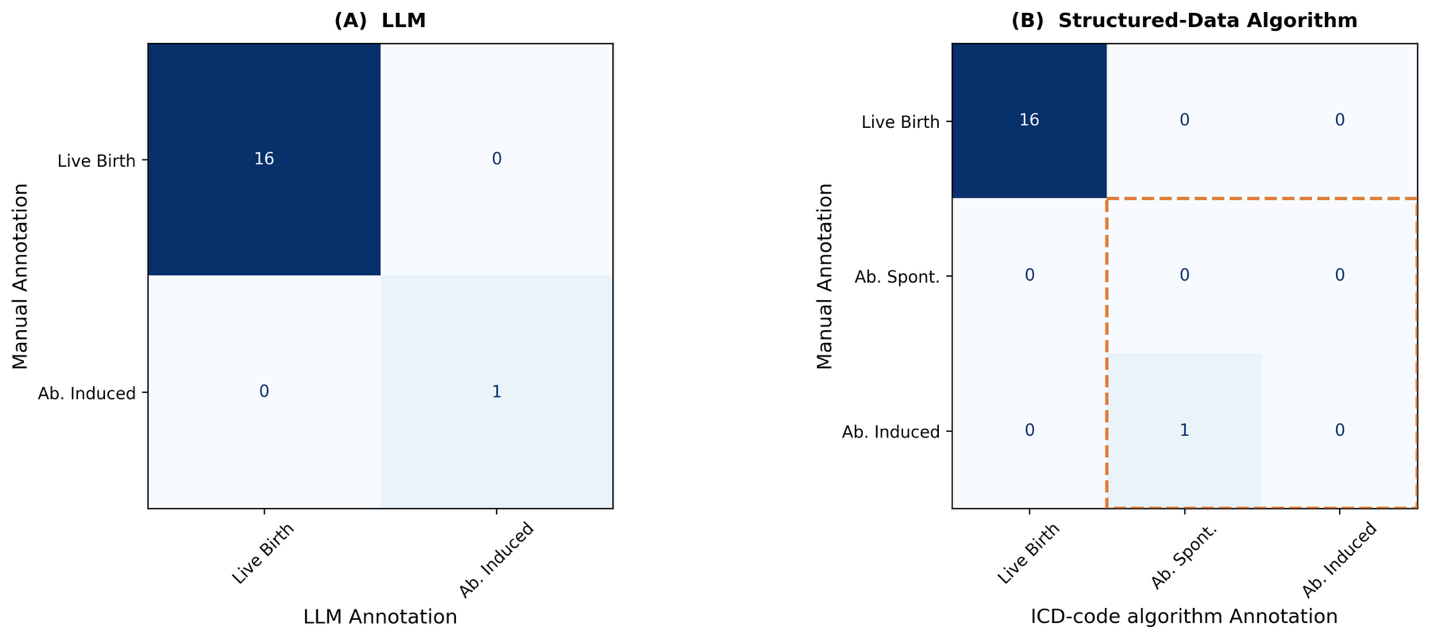

### Tables

#### Supplemental Table 1: ICD-10-CM Code Mapping Used in the Rule-based Pregnancy Episode Identification Algorithm

The table depicts which ICD codes were used for specific steps in the rule-based algorithm.

| **Category** | **Role in algorithm** | **ICD-10-CM codes** |
| --- | --- | --- |
| ***Encounter identification & gestational age*** | | |
| **Early pregnancy** | Flag early pregnancy encounters; back-calculate LMP (~10 wk) | 'Z32.01', 'Z34.01', 'Z34.81', 'Z34.91', 'O09.01', 'O09.11', 'O09.A1', 'O09.211', 'O09.291', 'O09.31', 'O09.41', 'O09.511', 'O09.521', 'O09.611', 'O09.621', 'O09.71', 'O09.811', 'O09.821', 'O09.891', 'O09.91','O30.001','O30.011','O30.021','O30.031','O30.041','O30.091', 'O20', |
| **Gestational age — exact (Z3A)** | Highest-priority GA source; derive LMP by subtraction | Z3A.04–Z3A.42 |
| **Gestational age — trimester last-character** | Assign midpoint 10/20/34 week when code suffix = 1/2/3 | 'O09','O10','O11','O12','O13','O14','O15','O16','O30','O34','O42','O43',  'O44','O45','O46','O47','O48' |
| **Gestational age — named trimester codes** | Assign trimester midpoint | first_trimester': ['Z34.01', 'Z34.81', 'Z34.91', 'O09.01', 'O09.11', 'O09.A1',  'O09.211', 'O09.291', 'O09.31', 'O09.41', 'O09.511', 'O09.521', 'O09.611', 'O09.621', 'O09.71', 'O09.811', 'O09.821', 'O09.891','O09.91','O30.001','O30.011','O30.021','O30.031','O30.041','O30.091','O99.351','O26.851','O99.611',  'O99.341','O99.411','O99.511','O99.811','O36.01','O36.11','O36.21','O36.31','O36.51','O36.61','O36.81','O36.91'],  'second_trimester': ['O09.02', 'O09.12', 'O09.A2', 'O09.212', 'O09.292', 'O09.32', 'O09.42', 'O09.512', 'O09.522', 'O09.612', 'O09.622', 'O09.72',  'O09.812', 'O09.822', 'O09.892', 'O09.92', 'Z34.02', 'Z34.82', 'Z34.92','O60.02','O30.002','O30.012','O30.022','O30.032','O30.042',  'O30.092','O99.352','O26.852','O99.612',  'O99.342','O99.412','O99.512','O99.812',  'O36.02','O36.12','O36.22','O36.32','O36.52','O36.62','O36.82','O36.92'],  'third_trimester': ['O09.03', 'O09.13', 'O09.A3', 'O09.213', 'O09.293', 'O09.33', 'O26.853','O99.613',  'O09.43', 'O09.513', 'O09.523', 'O09.613', 'O09.623', 'O09.73',  'O09.813', 'O09.823', 'O09.893', 'O09.93', 'Z34.03', 'Z34.83', 'Z34.93','O60.03','O30.003','O30.013','O30.023','O30.033','O30.043','O30.093',  'O99.353','O36.5930','O99.343','O99.413','O99.513','O99.813',  'O36.03','O36.13','O36.23','O36.33','O36.53','O36.63','O36.83','O36.93'] |
| ***Pregnancy outcomes (definitive outcome arm)*** | | |
| **Live birth** | Definitive outcome; triggers outcome-anchored episode arm | 'Z37.0', 'Z37.2', 'Z37.3', 'Z37.4', 'Z37.5', 'Z37.6', 'Z37.7', 'Z37.9', 'O80', 'O82','Z38' |
| **Stillbirth** | Definitive outcome | 'Z37.1', 'Z37.3', 'Z37.4','Z37.6', 'Z37.7' |
| **Preterm delivery** | Definitive outcome; GA midpoint assigned 36 week | 'O60.10', 'O60.12', 'O60.13','O60.14' |
| **Delivery** | Definitive outcome but undefined; 15-week post-outcome window | 'O81', 'O83', 'O84', 'O85','O63','O64','O65','O66','O67','O68','O69','O60.2' |
| **Spontaneous abortion / early loss** | Definitive outcome; 200-day lookback applied | 'O02','O03', 'O05', 'O06' |
| **Induced abortion** | Definitive outcome | 'O04', 'Z33.2' |
| **Ectopic pregnancy** | Definitive outcome; 120-day lookback; normal IUP codes excluded from episode | 'O00','O08' |
| **Molar pregnancy** | Definitive outcome | 'O01' |
| **Postpartum** | Grouped within 120-day window of delivery; outcome date offset −28 d; definitive outcome but undefined | 'Z39','O90','O89','O87','O86','O72', |
| ***Multiple gestation*** | | |
| **Twin pregnancy** | Set gestation_type = 'twin'; estimated_fetus_count = 2 | 'O30.0', 'O30.00', 'O30.001', 'O30.002', 'O30.003',  'O30.01', 'O30.011', 'O30.012', 'O30.013',  'O30.02', 'O30.021', 'O30.022', 'O30.023',  'O30.03', 'O30.031', 'O30.032', 'O30.033',  'O30.04', 'O30.041', 'O30.042', 'O30.043',  'O30.09', 'O30.091', 'O30.092', 'O30.093' |
| **Triplet pregnancy** | Set gestation_type = 'triplet'; estimated_fetus_count = 3 | 'O30.1', 'O30.10', 'O30.101', 'O30.102', 'O30.103',  'O30.11', 'O30.111', 'O30.112', 'O30.113',  'O30.12', 'O30.121', 'O30.122', 'O30.123',  'O30.19', 'O30.191', 'O30.192', 'O30.193' |
| **Other multiple gestation** | Set gestation_type = 'multiple_unspecified' | 'O30.2', 'O30.8', 'O30.9' |
| **Multiple gestation complications** | Flag is_multiple_gestation | 'O31' |
| ***Complications*** | | |
| **Preeclampsia / eclampsia** | Complication flag: has_preeclampsia | 'O14', 'O15', 'O11' |
| **Gestational hypertension** | Complication flag: has_gestational_hypertension | 'O13', 'O16' |
| **Gestational diabetes** | Complication flag: has_gestational_diabetes | 'O24', 'O24.4', 'O24.41', 'O24.42', 'O24.9' |
| **Hyperemesis gravidarum** | Complication flag: has_hyperemesis | 'O21', 'O21.0', 'O21.1', 'O21.2' |
| **Placental abruption** | Complication flag: has_placental_abruption | 'O45' |
| **Placenta praevia** | Complication flag: has_placenta_previa | O44 |
| **Chorioamnionitis** | Complication flag: has_chorioamnionitis | O41.12 |
| **Other placental abnormalities** | Complication flag: has_placental_abnormalities | 'O43', 'O44', 'O45', 'O46','O41.14' |
| **Fetal abnormalities** | Complication flag: has_fetal_abnormalities | O35, O26 |
| **Postpartum haemorrhage** | Complication flag: has_postpartum_hemorrhage | O72 |
| **Preterm labour** | Complication flag: has_preterm_labor | O60 |
| **Premature rupture of membranes** | Complication flag: has_premature_rupture_membranes | O42 |
| **Thromboembolic disorders** | Complication flag: has_thromboembolic_disorders | O22, O87 |
| **Infections** | Complication flag: has_infections | O98, O99.5 |
| **Obstructed labour** | Complication flag: has_obstructed_labor | O64–O66 |
| **Anesthesia complications** | Complication flag: has_anesthesia_complications | O29, O74, O89 |
| **Maternal sepsis** | Complication flag: has_maternal_sepsis | O85 |
| **Other obstetric complications** | Complication flag: has_other_obstetric_complications | O75, O94–O97 |
| **Postpartum complications (other)** | Complication flag: has_postpartum_complications | O85–O86, O90–O92 |

#### Supplemental Table 2: Matching of episodes in training data

The table shows per method the number of pregnancies that were successfully matched to human annotation, those that were missed (false negative), and additional ones (false positive) only identified by the LLM or structured data. A total of N=20 pregnancies were found in human annotation. Precision = TP/(TP+FP), recall = TP/(TP+FN), F1 = 2TP/(2TP+FP+FN), where TP are matched episodes.

| **Method** | **Matched** | **False Positive** | **False Negative** | **Precision** | **Recall** | **F1** |
| --- | --- | --- | --- | --- | --- | --- |
| LLM | 19 | 0 | 1 | 1.00 | 0.95 | 0.97 |
| Structured | 17 | 8 | 3 | 0.68 | 0.85 | 0.76 |

#### Supplemental Table 3: Train set performance for complications

The table displays for each complication the prevalence and performance of the two methods (LLM, structured data algorithm).

| **condition** | **source** | **sensitivity** | **specificity** | **f1** | **precision** | **n_pos** | **n** | **prevalence** |
| --- | --- | --- | --- | --- | --- | --- | --- | --- |
| chorioamnionitis | structured | 0 | 1 | 0 | 0 | 1 | 17 | 0.06 |
| chorioamnionitis | LLM | 0 | 1 | 0 | 0 | 1 | 17 | 0.06 |
| fetal_abnormalities | structured | 1 | 0.27 | 0.27 | 0.15 | 2 | 17 | 0.12 |
| fetal_abnormalities | LLM | 1 | 1 | 1 | 1 | 2 | 17 | 0.12 |
| gestational_diabetes | structured | 0 | 0.94 | 0 | 0 | 0 | 17 | 0 |
| gestational_diabetes | LLM | 0 | 1 | 0 | 0 | 0 | 17 | 0 |
| gestational_hypertension | structured | 0.8 | 0.83 | 0.73 | 0.67 | 5 | 17 | 0.29 |
| gestational_hypertension | LLM | 0.8 | 0.92 | 0.8 | 0.8 | 5 | 17 | 0.29 |
| hyperemesis | structured | 0 | 0.81 | 0 | 0 | 1 | 17 | 0.06 |
| hyperemesis | LLM | 1 | 1 | 1 | 1 | 1 | 17 | 0.06 |
| placenta_previa | structured | 0 | 1 | 0 | 0 | 0 | 17 | 0 |
| placenta_previa | LLM | 0 | 1 | 0 | 0 | 0 | 17 | 0 |
| placental_abnormalities | structured | 0.5 | 0.87 | 0.4 | 0.33 | 2 | 17 | 0.12 |
| placental_abnormalities | LLM | 1 | 1 | 1 | 1 | 2 | 17 | 0.12 |
| placental_abruption | structured | 0.5 | 1 | 0.67 | 1 | 2 | 17 | 0.12 |
| placental_abruption | LLM | 1 | 1 | 1 | 1 | 2 | 17 | 0.12 |
| postpartum_hemorrhage | structured | 1 | 0.86 | 0.75 | 0.6 | 3 | 17 | 0.18 |
| postpartum_hemorrhage | LLM | 1 | 0.93 | 0.86 | 0.75 | 3 | 17 | 0.18 |
| preeclampsia | structured | 1 | 1 | 1 | 1 | 1 | 17 | 0.06 |
| preeclampsia | LLM | 1 | 1 | 1 | 1 | 1 | 17 | 0.06 |
| Combined | structured | 0.71 | 0.86 | 0.48 | 0.36 | 17 | 170 | 0.1 |
| Combined | LLM | 0.88 | 0.99 | 0.88 | 0.88 | 17 | 170 | 0.1 |

#### Supplemental Table 4: Train set performance for pregnancy outcomes

The performance of the two methods (LLM, structured-data algorithm) are shown for assigning the correct outcome type to a pregnancy episode. The performance is given as a weighted score if it is multi-class, accuracy is not defined for these and thus left empty. The evaluation is once done on exact matching and once for a fuzzy matching where the individual abortion subtypes are merged into one broad category.

| **method** | **scoring** | **class_label** | **n_eval** | **accuracy** | **f1** | **precision** | **recall** |
| --- | --- | --- | --- | --- | --- | --- | --- |
| llm | exact | all | 17 | 1 | 1 | 1 | 1 |
| llm | fuzzy | live_birth | 17 | - | 1 | 1 | 1 |
| llm | fuzzy | abortion | 17 | - | 1 | 1 | 1 |
| llm | fuzzy | all | 17 | 1 | 1 | 1 | 1 |
| Structured | exact | all | 17 | 0.94 | 0.94 | 0.94 | 0.94 |
| Structured | fuzzy | live_birth | 17 | - | 1 | 1 | 1 |
| Structured | fuzzy | abortion | 17 | - | 1 | 1 | 1 |
| Structured | fuzzy | all | 17 | 1 | 1 | 1 | 1 |

#### Supplemental Table 5: Test set performance for pregnancy outcomes

The performance of the two methods (LLM, structured-data algorithm) are shown for assigning the correct outcome type to a pregnancy episode. The performance is given as a weighted score if it is multi-class, accuracy is not defined for these and thus left empty. The evaluation is once done on exact matching and once for a fuzzy matching where the individual abortion subtypes are merged into one broad category.

| **method** | **scoring** | **class_label** | **n_eval** | **accuracy** | **f1** | **precision** | **recall** |
| --- | --- | --- | --- | --- | --- | --- | --- |
| llm | exact | all | 38 | 0.84 | 0.87 | 0.90 | 0.84 |
| llm | fuzzy | live_birth | 38 | - | 0.94 | 1 | 0.89 |
| llm | fuzzy | abortion | 38 | - | 1 | 1 | 1 |
| llm | fuzzy | all | 38 | 0.92 | 0.95 | 0.97 | 0.92 |
| Structured | exact | all | 38 | 0.74 | 0.79 | 0.90 | 0.74 |
| Structured | fuzzy | live_birth | 38 | - | 0.91 | 1 | 0.84 |
| Structured | fuzzy | abortion | 38 | - | 0.94 | 1 | 0.89 |
| Structured | fuzzy | all | 38 | 0.84 | 0.90 | 0.97 | 0.84 |

#### Supplemental Table 6: Test set performance for complications

The table displays for each complication the prevalence and performance of the two methods (LLM, structured data algorithm).

| condition | source | sensitivity | specificity | f1 | precision | n_pos | n | prevalence |
| --- | --- | --- | --- | --- | --- | --- | --- | --- |
| chorioamnionitis | structured | 1 | 1 | 1 | 1 | 2 | 40 | 0.05 |
| chorioamnionitis | LLM | 0.5 | 1 | 0.67 | 1 | 2 | 40 | 0.05 |
| fetal_abnormalities | structured | 1 | 0.49 | 0.24 | 0.14 | 3 | 40 | 0.075 |
| fetal_abnormalities | LLM | 1 | 0.97 | 0.86 | 0.75 | 3 | 40 | 0.075 |
| gestational_diabetes | structured | 1 | 1 | 1 | 1 | 4 | 40 | 0.1 |
| gestational_diabetes | LLM | 1 | 1 | 1 | 1 | 4 | 40 | 0.1 |
| gestational_hypertension | structured | 0.67 | 0.95 | 0.57 | 0.5 | 3 | 40 | 0.075 |
| gestational_hypertension | LLM | 0.67 | 1 | 0.8 | 1 | 3 | 40 | 0.075 |
| hyperemesis | structured | 0 | 0.975 | 0 | 0 | 0 | 40 | 0 |
| hyperemesis | LLM | 0 | 1 | 0 | 0 | 0 | 40 | 0 |
| placenta_previa | structured | 0 | 1 | 0 | 0 | 0 | 40 | 0 |
| placenta_previa | LLM | 0 | 1 | 0 | 0 | 0 | 40 | 0 |
| placental_abnormalities | structured | 0.33 | 0.95 | 0.33 | 0.33 | 3 | 40 | 0.075 |
| placental_abnormalities | LLM | 0.67 | 1 | 0.8 | 1 | 3 | 40 | 0.075 |
| placental_abruption | structured | 1 | 1 | 1 | 1 | 1 | 40 | 0.025 |
| placental_abruption | LLM | 1 | 1 | 1 | 1 | 1 | 40 | 0.025 |
| postpartum_hemorrhage | structured | 0.5 | 0.97 | 0.5 | 0.5 | 2 | 40 | 0.05 |
| postpartum_hemorrhage | LLM | 1 | 0.95 | 0.67 | 0.5 | 2 | 40 | 0.05 |
| preeclampsia | structured | 1 | 0.97 | 0.8 | 0.67 | 2 | 40 | 0.05 |
| preeclampsia | LLM | 0.5 | 1 | 0.67 | 1 | 2 | 40 | 0.05 |
| Combined | structured | 0.8 | 0.93 | 0.52 | 0.38 | 20 | 400 | 0.05 |
| Combined | LLM | 0.8 | 0.99 | 0.82 | 0.84 | 20 | 400 | 0.05 |

#### Supplemental Table 7: Units of analysis and denominators

The table lists for each analysis the unit of analysis, the set of pregnancies it is based on, and the resulting N in the training and test sets. Pregnancy-level information was derived from the fetus-level annotation by taking the maximum across fetuses. “Matched by both methods” denotes annotated pregnancies for which both the LLM and the structured algorithm produced a matching episode; all comparisons between the two methods are restricted to this set. Pregnancies matched only by the LLM (train: 2, test: 13) or only by the structured algorithm (train: 0, test: 0) and unmatched predicted episodes (false positives) are therefore not part of these comparisons.

| **Analysis** | **Unit** | **Set** | **Train N** | **Test N** |
| --- | --- | --- | --- | --- |
| Cohort description (Table 1) | Patient | All annotated patients | 10 | 20 |
| Cohort description (Table 1) | Pregnancy | All annotated pregnancies | 20 | 54 |
| Cohort description (Table 1) | Fetus | All annotated fetuses | 21 | 55 |
| Episode detection, recall (Table 2, Supplemental Table 2) | Pregnancy | All annotated pregnancies | 20 | 54 |
| Episode detection, precision (Table 2, Supplemental Table 2) | Predicted pregnancy episode | All episodes predicted by each method | LLM 19; structured 25 | LLM 56; structured 64 |
| Start and end dates (Figure 1, Supplemental Figure 2) | Pregnancy | Matched by both methods, start and end dates available from all sources | 14 | 32 |
| Outcome type (Figure 2, Supplemental Tables 4-5) | Pregnancy | Matched by both methods, annotated outcome available | 17 | 38 |
| Complications (Figure 3, Supplemental Tables 3 and 6) | Pregnancy x complication | Matched by both methods | 17 (170 pairs) | 40 (400 pairs) |
| Assisted reproductive treatment, LLM vs structured | Pregnancy | Matched by both methods | 17 | 40 |
| Assisted reproductive treatment, LLM only | Pregnancy | Matched by the LLM | 19 | 53 |
| Multiple gestation | Pregnancy | Matched by both methods | 17 | 40 |
| Number of fetuses and fetus-level outcomes (LLM vs chart review) | Fetus | Fetuses of pregnancies matched by both methods | 18 (17 pregnancies) | 41 (40 pregnancies) |
